# Effects of Arylsulfatase B and Pembrolizumab in Combination on Progression of Metastatic Melanoma in the B16F10 Syngeneic Mouse Model

**DOI:** 10.64898/2026.01.02.25343295

**Authors:** Sumit Bhattacharyya, Insug O-Sullivan, Joanne Tobacman

## Abstract

In this report, experiments assessed how Arylsulfatase B (ARSB; N-acetylgalactosamine-4-sulfatase) treatment might interact with Pembrolizumab and improve therapeutic responses in melanoma. ARSB acts to remove 4-sulfate groups from chondroitin 4-sulfate (C4S; chondroitin sulfate A; CSA). ARSB is required for the degradation of C4S, identified as an oncofetal, tumor-agnostic antigen. Previous reports showed that in syngeneic B16F10 mouse melanomas, ARSB inhibited progression of subcutaneous and metastatic pulmonary melanomas and improved survival by direct effects on melanoma cells. ARSB enhanced apoptosis, which was mediated by increased expression of Constitutive Photomorphogenic (COP)1, an E3 ubiquitin ligase. Combined treatment by recombinant human ARSB, directed at melanoma cells, and Pembrolizumab, directed at infiltrating cytotoxic lymphocytes, can lead to increased apoptosis by different mechanisms, to declines in metalloproteinases and invasiveness, and to altered expression of cytokines. The apparent synergism between effects of ARSB and Pembrolizumab may further inhibit the progression of melanoma and improve treatment outcomes.

## INTRODUCTION

The enzyme ARSB (Arylsulfatase B; N-acetylgalactosamine-4-sulfatase) removes 4-sulfate groups from N-acetylgalactosamine-4-sulfate, specifically 4-sulfate groups at the non-reducing end of chondroitin 4-sulfate (C4S; chondroitin sulfate A; CSA) and dermatan sulfate (DS). Congenital, inherited inactivating mutations of ARSB lead to the diffuse accumulation of these sulfated glycosaminoglycans in the lysosomal storage disorder Mucopolysaccharidosis (MPS) VI. MPS VI is characterized by organ dysfunction, respiratory insufficiency, skeletal deformities with short stature and cartilage abnormalities, and reduced life expectancy [1]. Treatment with recombinant human (rh) ARSB is effective and well-tolerated, leading to significant clinical improvement [2]. In multiple reports, we have shown that acquired decline in ARSB function acts as a tumor promoter, including in melanoma, colon, prostate, and mammary tissues [3–11]. Malignancy results from the disruption of normal intra- and intercellular signaling due to increased chondroitin 4-sulfation and leads to transcriptional events. These effects are mediated by epigenetic mechanisms, as well as the activation of multiple transcription factors, including AP-1, Sp1, Smad3, c-Myc, and MITF, and by enhanced Wnt signaling [3,6,10–15]. C4S is also known as chondroitin sulfate A, which is recognized as an oncofetal antigen and the target of the malarial protein VAR2CSA [17–22].

The diverse signaling effects following decline in ARSB function are attributable to increased sulfation and accumulation of C4S when ARSB activity declines, leading to changes in the extent of binding of critical molecules with C4S. Critical effects include reduced binding of galectin-3 with more sulfated C4S and increased binding of SHP2 (PTPN11), the ubiquitous non-receptor tyrosine phosphatase, with more sulfated C4S. Increased binding of SHP2 with C4S leads to sustained phosphorylation of vital signaling molecules, including phospho-p38 MAPK and phospho-ERK1/2, and profound disruption of intracellular signaling [3,6,11–15]. These findings demonstrate that decline in ARSB function acts as a tumor promoter. Recent treatment studies with rhARSB in the B16F10 syngeneic melanoma model show that recombinant human ARSB acts as a tumor suppressor, associated with improved survival and with smaller and fewer subcutaneous tumors and pulmonary metastases [4–6].

These background findings reveal major effects of ARSB and C4S on melanoma cells, and suggest that combination with other therapeutic agents, such as immune checkpoint inhibitors (ICI) which enhance the attack on tumor cells by infiltrating immune cells, may be more effective than either rhARSB or ICI alone. Detailed analysis of the structural requirements for interaction between immune cell PD-1 and tumor PD-L1 has enabled development of effective antibodies to block this interaction, enhance cytotoxic immune responses, and provide clinical benefit [23–26]. With consideration of the potential clinical benefit of rhARSB treatment, either alone or in combination with checkpoint inhibition, studies were undertaken to examine how rhARSB and the anti-PD-1 agent Pembrolizumab interact in the treatment of metastatic pulmonary melanomas in the B16F10 syngeneic model and in cell-based experiments. Since treatment by ARSB reduces expression of PD-L1 [10], we anticipated that Pembrolizumab effectiveness might be enhanced in combination with ARSB. Experiments tested how ARSB and Pembrolizumab, alone and in combination, impacted on apoptosis, viability, invasiveness, cytokine/chemokine expression, and responses to peripheral blood mononuclear cells (PBMC) and polymorphonuclear leukocytes (PMN) in the syngeneic, murine B16F10 pulmonary melanoma model and in cell-based experiments in human melanoma cells.

## MATERIALS AND METHODS

### Syngeneic B16F10 pulmonary melanomas in C57BL/6J mice

Eight-week-old male C57BL/6J mice (n=32) were purchased (Jackson Laboratories, Bar Harbor, Maine) and housed in the Veterinary Medicine Unit at the Jesse Brown Veterans Affairs Medical Center (JBVAMC, Chicago, IL). Principles of laboratory animal care were observed, and all procedures were approved by the AALAC accredited Animal Care Committee of the JBVAMC. Mice were fed a standard diet and maintained in groups of four in a cage with routine light–dark cycles, as previously [4,5]. Mice were ear-punched for identification and divided into four groups, which were inoculated with 250,000 B16F10 tumor cells in saline by tail vein injection on day 0. B16 F10 cells (CRL-6475, ATCC, Manassas, VA; RRID: CVCL_0159) were cultured in DMEM (ATCC 30-2002) with 10% FBS under the recommended conditions. Recombinant bioactive ARSB (CF 4415-SU, R&D Systems, Bio-Techne, Minneapolis, MN) at a dose of 0.2 mg/kg BW in normal saline was injected into the tail vein. Eight untreated mice received parallel vehicle (saline) injections. Pembrolizumab was purchased (HY-P9902, MedChemExpress, Mammoth Junction, NJ) and injected intraperitoneally at a concentration of 5 mg/kg. Treatment groups included: rhARSB-treated (0.2 mg/kg IV on days 2, 7, 12); Pembrolizumab (5 mg/kg IP on days 2, 7, and 12); and combined Pembrolizumab (5 mg/kg IP and rhARSB IV on days 2, 7, 12). Two mice died spontaneously of uncertain cause prior to day 14, including an untreated mouse on day 2 and a mouse with combined treatment on day 12. 30 mice were euthanized on day 14 by isoflurane inhalation and decapitation. At the time of euthanasia, lung, liver, spleen, blood, and other organs were harvested. Pulmonary nodules were counted by two observers and photographed.

### Cell culture of A375 melanoma cells and normal human melanocytes

A375 human melanoma cells (CRL-1619, ATCC, Manassas, VA; RRID: CVCL_0132) were grown in Dulbecco’s modified Eagle medium (DMEM) with 10% fetal bovine serum (FBS; ATCC) and 1x penicillin-streptomycin (ATCC). Cells were maintained at 37°C in a humidified, 5% CO_2_ environment with media exchange every 2 days. Confluent cells in T-25 flasks were harvested using EDTA-trypsin (ATCC) and sub-cultured. Cells were screened for pathogens by IDEXX BioAnalytics (Columbia, MO).

### Silencing of insulin-like growth factor 2 receptor (IGF2R), N-acetylgalactosamine-4-sulfatase (Arylsulfatase B, ARSB) and Interleukin (IL)-8 by siRNA

The mRNA expression of IGF2R and of ARSB was silenced by specific siRNA using previously reported procedures [4–6,13]. Small interfering (si) RNA for IGF2R (Assay ID s7218, Thermo Fisher Scientific, Waltham, MA) and siRNA for ARSB siRNA were previously detailed [13,15]. Validated IL-8 (CXCL8) siRNA (Assay ID s7327, Thermo Fisher) was used with similar procedures. A375 cells were grown to ∼70 % confluence, then silenced by adding 0.6 μl of 20 μM siRNA (150 ng), mixed with 100 μl of serum-free medium and 12 μl of HiPerfect Transfection Reagent (Qiagen, Germantown, MD). Media were changed after 24 h, and cell treatments were initiated.

### Pembrolizumab binding assay

To assess effectiveness of the human PD-1 antibody Pembrolizumab in our syngeneic mouse model, the binding of the mouse recombinant PD-1 protein with hFc Tag (Catalog #: 50124-M02H, SinoBiological, Houston, TX) to Pembrolizumab-coated wells in a microplate was measured and compared with isotype IGGĸ4 control. Pembrolizumab 100 µl (4 µg/ml, MedchemExpress, Monmouth Junction, NJ; HY-P99002) or isotype control (IGG4-kappa; HY-P99003) in PBS was used to coat wells of a 96-well microplate, and the plate was incubated overnight at 4°C. Subsequently wells were washed and blocked with 300 µl of blocking buffer for 2h at RT, washed x3 and mouse PD-1 protein with human Fc tag was added in diluent and incubated overnight at 4°C. Wells were washed and 100 µl of the secondary HRP-antibody to the human Fc tag (Catalog #: NB7449, Novus Biologicals, Centennial, CO) at a dilution of 1:10,000 was added for 90 minutes at RT. Wells were washed and 100 µl of substrate (1:1 mixture of Color Reagent A (H_2_O_2_) and Color Reagent B (tetramethylbenzidine) were added. and the plate was incubated in the dark at RT for of mouse PD-1 and the extent of isotype control binding was read at 450 nm. Isotype control binding to mouse PD-1 was minimal, in contrast to binding to Pembrolizumab-coated wells (Supplementary Fig.5A). Binding to Pembrolizumab increased with increasing concentration of mouse PD-1, peaking at ∼1.0 µg/ml (Supplementary Fig.5B).

### Resazurin-based cell viability assay

Viability assay was performed using the PrestoBlue Cell Viability reagent (Catalog #: A13261; ThermoFisher). A365 and B16F10 melanoma cells (25,000 cells/well) were plated in wells of a microplate in their growth media. After 24 hours, media were exchanged and 25,000 activated PBMC, Pembrolizumab (2 µg/ml), or rhARSB (1 ng/ml), or the combinations of rhARSB and activated PBMC, Pembrolizumab and activated PBMC, or rhARSB, Pembrolizumab and activated PBMC were added to 6 wells (total of 36 wells and 6 control wells for each cell line). After 24 hours, media were exchanged, cells washed, and fresh media and PrestoBlue were added. After 10 minutes at RT, absorbance was detected and compared among the groups.

### Treatment of A375 human melanoma and melanocytes cells by exogenous ARSB and other agents

Cells were treated by exogenous, bioactive rhARSB (1 ng/ml x 24 h; R&D, Bio-Techne) for 24 h, unless indicated otherwise. Cells were treated with ERK activation inhibitor peptide 1, cell-permeable (10 μM; Sigma-Aldrich, Inc., St. Louis, MO); F4, a cell-permeable thiazolidinone compound which inhibits c-Myc-Max dimerization (64 μM, 10058-F4, Tocris Bioscience, Bio-Techne); and p38 MAPK inhibitor SB203580 (10 µM; Tocris Bioscience, Bio-Techne). Some preparations were treated with recombinant, human IL-8 (CXCL8 (1 ng/ml x 24h, 208-1L, R&D Systems).

### ELISAs for cleaved caspase-3 (Asp175), IL-8, MMP2, and Granzyme B

Cleaved caspase 3 in the cell extracts was measured by a commercial sandwich duoset ELISA (DYC835, R&D Systems), following the instructions. Samples and standards were added to the wells of a microtiter plate precoated with a capture antibody. Cleaved caspase 3 in the cell extracts was captured by the coated antibody on the plate and detected with a biotinylated second antibody to cleaved caspase 3. Streptavidin-HRP and hydrogen peroxide / tetramethylbenzidine (TMB) substrate were used to develop color proportional to the bound HRP activity. Similar ELISA procedures were used to quantify mouse matrix metalloproteinase (MMP)2 (ELM-MMP2, RayBiotech, Peachtree Corners, GA, USA), IL-8 (DY208, R&D Systems), IL-8 (DY208, R&D Systems), and Granzyme B (DY2906, R&D Systems). Results are reported as pg/mg protein, pg/ml or % control.

### QRT-PCR for COP1, BCL2, MMP2, MMP9, MCP1, IL-10, IL-17α, KC, IL-6, TNFα, bFGF, VEGF, and EGF

Total RNA was prepared from treated and control cells or tissue using an RNeasy Mini Kit (Qiagen). Equal amounts of purified RNAs from the control and treated preparations were reverse-transcribed and amplified using Brilliant SYBR Green QRT-PCR Master Mix (Bio-Rad, Hercules, CA). Human β-actin was used as an internal control. QRT-PCR was performed using the following specific primers (IDT, Integrated DNA Technologies, Coralville, IA):

Human β-actin (NM_001101) left: 5’-CACCATTGGCAATGAGCGGTTC-3’ and right: 5’-AGGTCTTTGCGGATGTCCACGT-3’;
Human BCL2 (NM_000633) left 5′-ATCGCCCTGTGGATGACTGAGT-3′ and right 5′-GCCAGGAGAAATCAAACAGAGGC-3′;
Human COP1 (NM_022457) left 5′-GTCAGTGAGGATAGCACAGTGC-3’ and right 5′-GAGAACTGCCACTGAAACCTGG-3’;
Human EGF (NM_001963) left 5’- TGCGATGCCAAGCAGTCTGTGA-3’ and right 5’-GCATAGCCCAATCTGAGAACCAC;
Human FGF2 (NM_002006) left 5’- AGCGGCTGTACTGCAAAAACGG-3’ and right 5’-CCTTTGATAGACACAACTCCTCTC;
Human IL-6 (NM_000600) left: 5’-AAAGCAGCAAAGAGGCACTG-3’ and right 5’-TTTTCACCAGGCAAGTCTCC-3’;
Human IL-8 (NM_00584) left5’-GAGAGTGATTGAGAGTGGACCAC-3’ and right 5’-CACAACCCTCTGCACCCAGTTT-3’;
Human IL10 (NM_000572) left 5’-TCTCCGAGATGCCTTCAGCAGA-3’ and right 5’-TCAGACAAGGCTTGGCAACCCA;
Human IL-17a (NM_002190) left 5’- CGGACTGTGATGGTCAACCTGA-3’ and right 5’-GCACTTTGCCTCCCAGATCACA;
Human MCP1 (NM_002982) left 5’-GAGATCACCAGCAGCAAGGTGCC-3’ and right 5’-TCCTGAACCCACTTCTGCTTGG-3’;
Human MMP2 (NM_004530) left: 5′-AGTGGATGATGCCTTTGCTC-3′ and right: 5′-GAGTCCGTCCTTACCGTCAA-3′;
Human MMP9 (NM_004994.2) left: 5′-GTCTTCCCCTTCACTTTCCTG-3′ and right: 5′-TCAGTGAAGCG GTACATAGGG-3′;
Human TNFα (NM_000594) left 5’- CTCTTCTGCCTGCTGCACTTTG and right 5’-ATGGGCTACAGGCTTGTCACTC
Human VEGFA (NM_001025366) left 5’- TTGCCTTGCTGCTCTACCTCCA and right 5’-GATGGCAGTAGCTGCGCTGATA-3’;
Mouse β-actin (NM_007393) left: 5‘-CATTGCTGACAGGATGCAGAAGG-3’ and right: 5’-TGCTGGAAGGTGGACAGTGAGG-3’;
Mouse BCL2 (NM_009741) left: 5′-CCTGTGGATTGAGTACCTG-3′ and right 5′-AGCCAGGAGAAATCAAACAGAGG-3′;
Mouse COP1 (NM_011931) left: 5′-GCCTCTACTCTCTCAGTGA -3′ and right: 5′-GTCCCACTGAAACCTGGAGGTT-3′;
Mouse EGF (NM_010415) left 5’- GAGTTCCGTACTCCCTCTTGCA and right 5’-CAGCCAAGACTGTAGTGTGGTC-3’;
Mouse bFGF (NM_008006) left 5’- AAGCGGCTCTACTGCAAGAACG-3’ and right 5’-CCTTGATAGACACAACTCCTCTC-3’;
Mouse IL6 (NM_031168) left 5’-AGTTGCCTTCTTGGGACTGA-3’ and right 5’-TCCACGATTTCCCAGAGAAC-3’;
Mouse IL-10 (NM_010548) left 5’-CGGGAAGACAATAACTGCACCC-3’ and right 5’-CGGTTAGCAGTATGTTGTCCAGC-3’;
Mouse IL-17a (NM_010552) left 5’- CAGACTACCTCAACCGTTCCAC-3’ and right 5’-TCCAGCTTTCCCTCCGCATTGA-3’;
Mouse KC (NM_008176) left 5’- TCCAGAGCTTGAAGGTGTTGCC-3’ and right 5’-AACCAAGGGAGCTTCAGGGTC-3’;
Mouse MCP1 (NM_011333.3) left 5’-ATCTGCCCTAAGGTCTTCAGC-3’ and right 5’-TAAGGCATCACAGTCCGAGTC-3’.
Mouse MMP2 (NM_008610) left 5’-CAGTGATGGCTTCTTGTGGT-3’ and right 5’-GGTCATAGTCCTCGGTGGTG-3’;
Mouse MMP9 (NM_0113599.5) left 5’-GACTACGATAAGGACGGCAAA-3’ and right 5’-CTCAAAGATGAACGGGAACAC-3’;
Mouse VEGF (NM_001025250) left 5’- for CTGCTGTAACGATGAAGCCCTG-3’ and right 5’-GCTGTAGGAAGCTCATCTCTCC-3’.

Cycle threshold (Ct) was determined during the exponential phase of amplification, as previously [15]. Fold changes in expression were determined by the difference between the Ct values of the gene of interest and the control.

### Transcription Factor filter plate assay for c-Myc by DNA hybridization

Transcription Factor (TF) Filter Plate Assay kit and specific labeled DNA probe for c-Myc were obtained (Signosis, Santa Clara, CA). Nuclear extracts were obtained from treated and control cells and were mixed with a specific biotin-labeled c-Myc DNA binding sequence and TF-DNA complexes formed. The assay filter plate retained the bound DNA probe and free probe was removed. The bound, pre-labeled DNA probe was then eluted from the filter and collected for detection by streptavidin-HRP and quantitative analysis. Luminescence was reported as relative light units (RLUs) on a microplate reader (FLUOstar, BMG Labtech, Cary, NC). The luminescence was normalized by total protein of the nuclear extract and expressed as percentage of control.

### A375 migration assay

Invasiveness of A375 human melanoma cells was detected using a fluorescent cell invasion assay (QCM^TM^ ECMatrix cell invasion assay (ECM550, Millipore Sigma, Burlington, MA). A 96-well cell culture plate and cell culture inserts with 8 μm pores in a polycarbonate membrane and coated with a thin, dried layer of ECMatrix^TM^ to block non-invasive cell migration, were used. A375 melanoma cells migrated through the ECM layer and attached to the bottom of the polycarbonate membrane. The cells were dissociated from the membrane by incubation with Cell Detachment Buffer. Then, the migrated cells were lysed and detected by a green-fluorescent dye (CyQuant GR dye, Thermo Fisher Scientific) using a fluorescence plate reader (BMG Labtech)..

### Polymorphonuclear (PMN) Leukocyte invasion assay

Human polymorphonuclear (PMN) cells (SER-PMN, PMN 042324B Blue 34/4 #65) were procured (Zenbio, Durham, NC). 350 µl of frozen cells containing about 5.25 x 10^6^ million cells were thawed and washed twice with normal saline and then resuspended in 2.5 ml DMEM containing 10% FBS. Calcein-AM (5 μg/ml; Sigma-Aldrich) was added to the 2.5 ml suspension of cells in DMEM-FBS, and the cells were incubated for 30 min at 37°C. PMN were washed twice with normal saline, counted in a counting chamber, and resuspended in DMEM-FBS to the desired concentration of 2,000 cells/µl. To assess migration of the PMN to melanoma cells, 200,000 A375 cells were seeded into twenty-four well plates and treated with either rhARSB or silencing RNAs (ARSB and control) for 24 h. Disposable 24-well transwell filter inserts were placed inside each well (Corning, Thermo Fisher #07-200-174). The inserts for the 24-well plate were 6.5 mm in diameter, with culture area of 0.33 cm^2^, and with pore size of 8 μm. 100 µl PMN suspensions were added directly onto the filters in each well, and the plate was incubated for 1 h (37°C and 5% CO_2_). The non-migrating cells on the top of the filter were removed gently. The cells that migrated into the bottom chamber were detected and measured by using the calcein fluorescence signal (excitation at 485 nm; emission at 530 nm) in a fluorescent plate reader (BMG Labtech).

### Co-culture of PBMCs and PMNs with A375 Melanoma cells

A total of 10^5^ A375 melanoma cells/well were seeded into twenty-four well plates in routine growth media, and siRNA for ARSB or control was added to some of the wells. After 24 h, media was removed and replenished with fresh media with or without ARSB (1 ng/ml). In some wells, 10^5^ PBMCs (PCS-800-011, ATCC), activated or non-activated, 10^5^ PMN, or Pembrolizumab 2 ug/ml, either alone or in combination with other treatments, was added. PBMC were activated by treatment with anti-CD3 (10 µg; 16-0037-81, Invitrogen, Thermo Fisher; RRID:AB_468854) and anti-CD28 (10 µg, 16-0281-81, Invitrogen; RRID:AB_46892) to 10^6^ PBMCs in10 ml of melanoma media in a T-25 flask. The cells were incubated with the antibodies at 37°C for 2 h for activation, and then were incubated for 24 h at 37°C and 5% CO_2_. Then, the media was removed and saved, and the cells were washed with DPBS, and harvested by scraping.

### Detection of IL-8 bound to chondroitin 4-sulfate

96-well ELISA plates were coated with 5 µg/ml of chondroitin 4-sulfate monoclonal antibody (Clone LY111, TCI, Portland, OR) or chondroitin 6-sulfate antibody (Clone MC21C, LSBio, Neward, CA) in PBS by adding 100 µl of antibody solution to each well and incubating at room temperature (RT) overnight. After antibody coating, the wells were washed three times with wash buffer (PBS 7.4 with 0.05% Tween 20) and blocked for 1 h at RT with Blocking Buffer (PBS 7.4 with 1% BSA). After blocking, 100 µl of chondroitin sulfate C (C6S) (2.5 µg/ml) from shark cartilage (predominantly C6S; Seikagaku, #400670, Tokyo, Japan; 73% 6S, 19 ± 1% 4S, 8% 2S6S, and 1% unsulfated chondroitin) or chondroitin sulfate A (C4S) (2.5 µg/ml) from bovine trachea (predominantly C4S; Sigma C9819; 86 ± 1% C4S, 12 ± 1% C6S, and 2% unsulfated chondroitin) was added to designated wells and incubated at 4°C overnight. Wells were washed three times with wash buffer, and 100 µl of recombinant human Interleukin-8 (208-IL, OriGene Technologies, Rockville, MD) at varying concentrations (20-1280 pg/ml) was added to specified wells and incubated at RT for 2 h. Wells were washed, 100 µl of biotinylated IL-8 antibody (DY208, R&D) was added, and the plate was incubated at RT for 2 h. Subsequently, wells were washed three times, 100 µl of 1:200 Streptavidin-HRP was added in each well, and the plate was incubated at RT for 20 min. The color was developed by adding 100 µl of color reagent (1:1, H_2_O_2_:TMB), incubated for 15-20 min, and then the reaction was stopped by adding 50 µl of Stop Solution (2N H_2_SO_4_). The optical density was measured at 450 nm in an ELISA plate reader (BMG Labtech).

### MMP activity assay

Matrix metalloproteinase (MMP) activity was measured in the spent media of A375 cells using the MMP activity assay (ab112146, Abcam, Cambridge, MA) in which a fluorescence resonance energy transfer (FRET) peptide is the MMP substrate. In the intact FRET peptide, the endogenous fluorescence of one part of the peptide is quenched by another part of the peptide. Following cleavage into two fragments by MMP, the fluorescence is recovered and can be detected in the fluorescence microplate reader (BMG Labtech) at Excitation/Emission = 490/525 nm. A375 melanoma cells were grown in 24-well clusters, and the FRET peptide was added when cells were ∼70% confluent. Samples were incubated with an equal volume of 1 mM APMA (4-aminophenylmercuric acetate) at 37°C for 1 h (for MMP2) and green fluorescence measured.

### Statistical analysis

The data presented in the Figures and Supplementary Figures are the mean ± SD of at least six independent experiments, unless indicated otherwise. Statistical significance was determined by one-way ANOVA followed by Tukey post-test for multiple comparisons. Analysis was performed using Prism (Version 10.3, GraphPad, CA) software. In the figures, **** represents p≤0.0001, *** for p≤0.001, ** for p≤0.01, and * for p≤0.05. The data generated in this study are available from the corresponding author (JKT) upon request.

## RESULTS

### Recombinant human (rh) ARSB and Pembrolizumab inhibit progression of metastatic pulmonary melanomas in the B16F10 syngeneic mouse model

Four groups of 10.5-week-old male C57BL/6J mice were inoculated with B16F10 melanoma cells to produce pulmonary melanoma metastases (**Fig.1A-1D**). One group (n=8) was treated by tail-vein injection of rhARSB 0.2 mg/kg (concentration of 50 µg/ml in normal saline) on days 2, 7, and 12 following tumor inoculation. A second-group (n=8) was treated by the anti-PD-1 checkpoint inhibitor Pembrolizumab (5 mg/kg IP) on days 2, 7, and 12 following tumor inoculation. A third group (n=7) was treated by the combination of rhARSB (0.2 mg/kg IV) and Pembrolizumab (5 mg/kg IP) on days 2, 7, and 12. A control group (n=7) was untreated and received normal saline IV on days 2, 7, and 12 (**Fig.1A**). Comparison of the lung images indicates that the untreated mice had many more metastatic tumors than the treated mice. Tumors were quantified by unblinded observers, and the rhARSB-treated mice had the fewest tumors (**Fig.1E**). All treatment groups had fewer metastases than the saline-treated controls. One untreated mouse had macroscopic liver metastases, and one control and one combined treatment mouse died spontaneously of uncertain cause several days prior to termination of the study on day 14. These findings are consistent with previous reports that treatment by rhARSB reduced the size and increased the survival of mice with B16F10 subcutaneous melanomas [5] and reduced the number of pulmonary metastases in female mice with B16F10 syngeneic melanomas [4]. Since the homology of the extracellular domain of human PD-1 and mouse PD-1 is only 62%, the full impact of checkpoint inhibitor treatment is not demonstrable in the syngeneic mouse model [23].

**Fig. 1.**
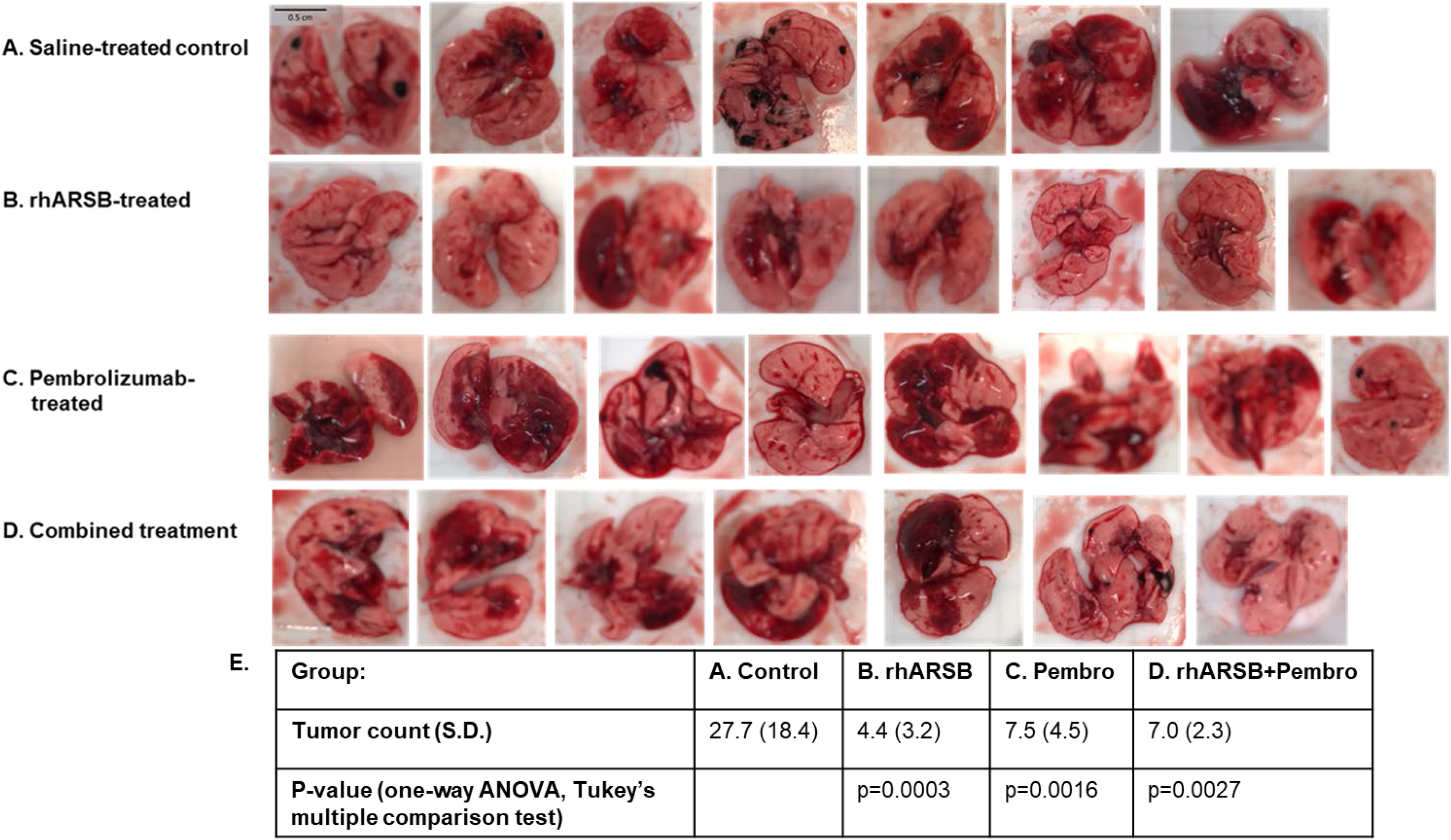
Impact of ARSB and Pembrolizumab and their combination on pulmonary metastases in B16F10 melanoma. Thirty-two 10.5 week-old male C57BL/6J mice were inoculated with 250,000 B16F10 melanoma cells by tail vein injection. Two mice died of uncertain causes, and the remaining mice were euthanized on day 14 following tumor inoculation. Groups were: **A.** untreated; **B.** rhARSB (0.2 mg/kg IV on days 2, 7, and 12); **C.** Pembrolizumab (5 mg/ml IP on days 2,7, and 12); and **D.** combined rhARSB and Pembrolizumab treatments. **E**. Compared to saline-treated control, the rhARSB, Pembrolizumab, and the combined rhARSB and Pembrolizumab groups had significantly fewer tumors (one-way ANOVA with Tukey post-test for multiple comparisons). Bar = 0.5 cm. (ARSB=arylsulfatase B=N-acetylgalactosamine-4-sulfatase; Pembro=Pembrolizumab rh=recombinant human).

Additional experiments show that treatment by rhARSB inhibited development of A375 subcutaneous tumors in humanized NSG mice with PBMC implants (**Supplementary Fig. 1A**) and reduced the growth of B16F10 subcutaneous melanomas which were measureable prior to initiation of treatment (**Supplementary Fig. 1B**). No harmful effects of rhARSB or Pembrolizumab were observed in any of the treated mice. These results show that treatment by rhARSB alone acts to suppress tumor development in syngeneic and humanized mouse melanoma models and can retard the progression of existing melanomas.

### Combined effects of rhARSB and Pembrolizumab

Decline in pulmonary melanoma metastases following rhARSB was attributed to apoptosis, mediated by the E3 ubiquitin ligase constitutive photomorphogenic 1 (COP1) and the associated decline in BCL2 [4]. Here, combined treatment by rhARSB and Pembrolizumab produced greater increase in cleaved caspase-3 than either agent alone. (**Fig.2A**). COP1 expression increased following rhARSB treatment, but Pembrolizumab had no effect on COP1 expression (**Fig.2B**). Both rhARSB and Pembrolizumab reduced Bcl2 expression, and their combination further reduced Bcl2 in the pulmonary melanomas (**Fig.2C**). The Spearman correlation coefficient between Bcl2 and cleaved caspase-3 is -0.97, reflecting the impact of both rhARSB and Pembrolizumab on decline in Bcl2. Spearman correlation coefficient of COP1 and cleaved caspase-3 is 0.85, reflecting the impact of rhARSB-induced increase in COP1 on these measurements.

**Fig. 2.**
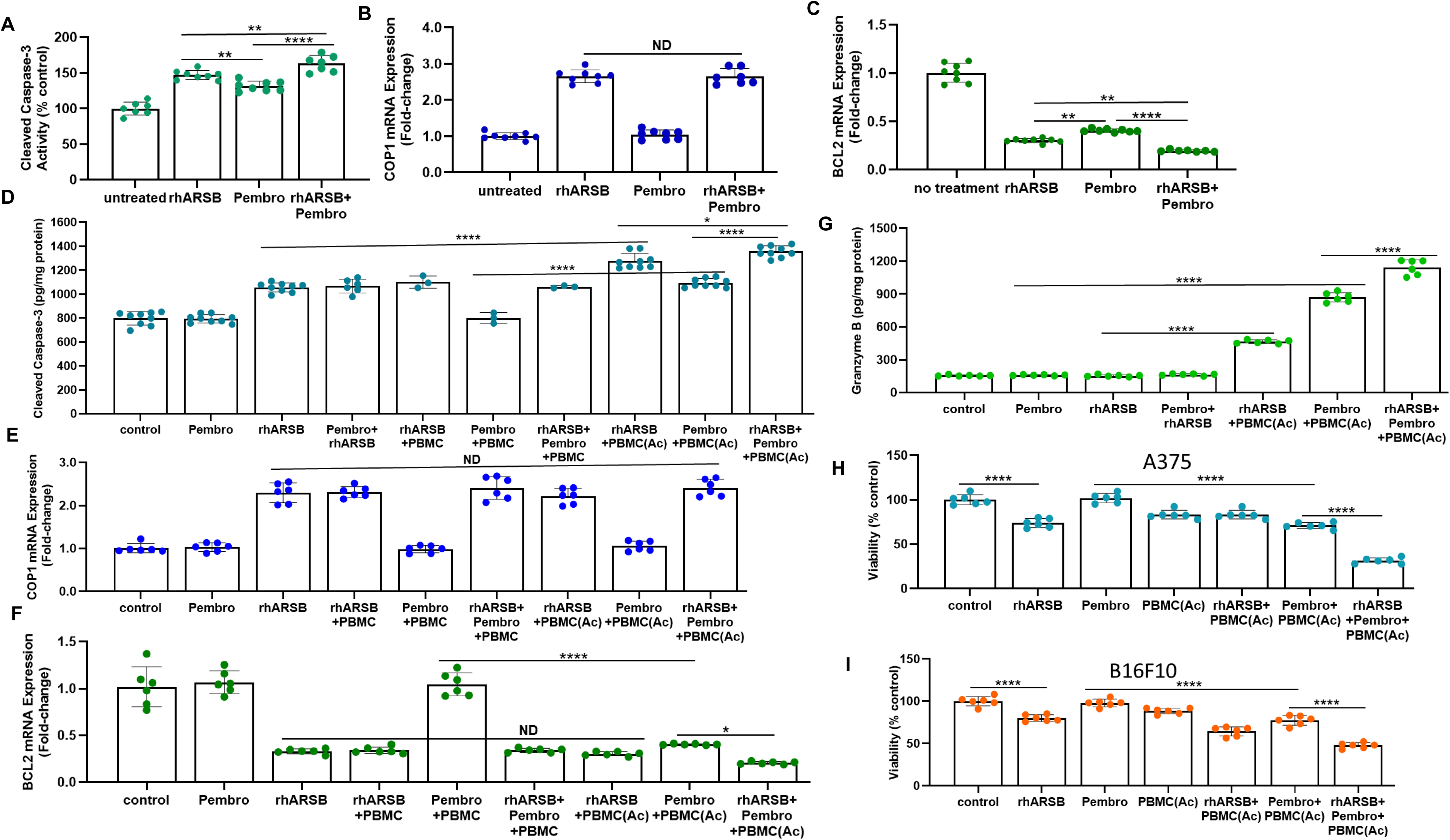
Effects of Pembrolizumab, ARSB, and their combination on COP1-mediated pathway of apoptosis in B16F10 pulmonary melanomas and A375 human melanoma cells. **A.** Cleaved caspase-3 increased following rhARSB or Pembrolizumab and was further Increased by their combination in treated B 16F10 pulmonary melanomas. All p-values are calculated by one-way ANOVA with Tukey test for multiple comparisons. **B.** In treated B16F10 melanomas, COP1 mRNA expression increased following rhARSB, but not following Pembrolizumab treatment. **C.** BCL2 mRNA expression declined following either rhARSB or Pembrolizumab and declined further by their combination in the B16F10 pulmonary melanomas. **D.** In cultured human A375 melanoma cells, cleaved caspase-3 increased following rhARSB and was further increased by the combination of rhARSB with activated PBMC [PBMC(Ac)]. Pembrolizumab alone or Pembrolizumab with unactivated PBMC did not increase cleaved caspase-3. However, cleaved caspase 3 was increased by the combination of Pembrolizumab with PBMC(Ac) and further increased by rhARSB, Pembrolizumab, and activated PBMC. **E.** In the A375 cells, COP1 increased following rhARSB, but was unaffected by Pembrolizumab or by PBMC, either activated or not activated. **F.** Inverse to the effect on COP1, rhARSB reduced the expression of BCL2. Neither Pembrolizumab alone nor PBMC alone affected BCL2 expression, but Pembro with activated PBMC reduced BCL2. BCL2 was further reduced by the combination of rhARSB + Pembrolizumab + PBMC(Ac) in the A375 cells. **G.** Granzyme B increased following the combinations of rhARSB and PBMC(Ac) and Pembrolizumab and PBMC(Ac). Synergistic effect was evident following treatment by rhARSB + Pembrolizumab + and PBMC(Ac) in the A375 cells. **H.** Viability was detected by the PrestoBlue assay in A375 cells. RhARSB, activated PBMC, and Pembrolizumab with activated PBMC and further increased by their combination. **I.** Viability was also reduced in the B16F10 cells following rhARSB, Pembrolizumab with activated PBMC, and activated PBMC and most reduced by the combination. Declines were less than in the A375 cells. [(Ac)=activated; PBMC(Ac)=activated peripheral blood mononuclear cells; COP1=constitutive photomorphogenic protein 1; Pembro=Pembrolizumab; PBMC=peripheral blood mononuclear cells; rhARSB=recombinant human arylsulfatase B]

In cultured A375 human melanoma cells treated with rhARSB, Pembrolizumab, peripheral blood mononuclear cells (PBMC), activated PBMC [PBMC(Ac)], and their combinations, cleaved caspase-3 increased following rhARSB or Pembrolizumab with activated PBMC, but not by Pembrolizumab alone or with unactivated PBMC (**Fig.2D**). Cleaved caspase-3 increased more by rhARSB with activated PBMC than by rhARSB alone, and increased most by the combination of rhARSB, Pembrolizumab, and activated PMBC. Recombinant human ARSB increased COP1 expression, and COP1 expression was not increased by Pembrolizumab or increased further by the combination of rhARSB or Pembrolizumab and activated PBMC (**Fig.2E**). These findings demonstrate a requirement for activated PBMC for Pembrolizumab-induced apoptosis and confirm the lack of impact of Pembrolizumab on COP1 expression shown in the mouse pulmonary melanomas.

In contrast to the lack of impact following Pembrolizumab and activated PBMC on COP1, rhARSB alone and Pembrolizumab with activated PBMC reduced BCL2 expression. The combination of rhARSB, Pembrolizumab and activated PBMC reduced BCL2 mRNA expression by about 80% compared to control (**Fig.2F**). Both rhARSB and Pembrolizumab with activated PBMC increased granzyme in the pulmonary melanoma tissue. Granzyme B increases apoptosis by BCL2-associated effects, and the increases in granzyme B reflect the involvement of antitumor, infiltrating immune cells in Pembrolizumab-induced apoptosis in melanoma [29–33]. The combination of rhARSB, Pembrolizumab, and activated PBMC showed a synergistic increase in granzyme B (**Fig.2G**).

In addition to effects on apoptosis, rhARSB, Pembrolizumab with activated PBMC, and the combination of rhARSB, Pembrolizumab, and activated PBMC significantly reduced the viability of A375 (Fig.2H) and B16F10 (Fig.2I) melanoma cells. Less reduction of resazurin to resorufin followed these treatments, consistent with decline in cell viability.

These experiments indicate distinct mechanisms of BCL2 inhibition by rhARSB, mediated by COP1 and by granzyme-induced effects in the presence of activated PBMC. Pembrolizumab had no effect on COP1, but had a greater effect on granzyme in the presence of activated PBMC than rhARSB and activated PBMC. The increase in cleaved caspase-3 activity and the increased reduction in BCL2 expression by the combination of rhARSB with Pembrolizumab *in vivo* or with Pembrolizumab and activated PBMC *in vitro* suggest a potential synergistic treatment benefit of these therapies in combination due to different mechanisms. Since COP1 is not modified by Pembrolizumab and activated PBMC, both the rhARSB-mediated increase in COP1 and the rhARSB, Pembrolizumab, and PBMC mediated increase in granzyme can contribute to apoptosis by decline in BCL2 and increase in cleaved caspase-3.

These results indicate that the tumor cell-based inhibitory effects of rhARSB combine with the immune-cell based effects of Pembrolizumab to augment apoptosis and reduce viability. The enhanced effects on increases in cleaved caspase-3 and granzyme B and, declines in BCL2 and viability suggest the potential for increased *in vivo* therapeutic effect,

### Exogenous ARSB reduces expression of matrix metalloproteinases (MMP) 2 and 9

In addition to effects of rhARSB on apoptosis in melanoma, rhARSB also reduces the expression of matrix metalloproteinases 2 and 9, two Type IV gelatinases which remodel the extracellular matrix and facilitate spread of malignant cells. Previously, decline in ARSB expression was shown to increase MMP2 and MMP9 [3,5,6,34,35]. In the pulmonary melanoma metastases, MMP2 and MMP9 expression declined significantly following rhARSB (**Fig.3A,3B**). Pembrolizumab had no effect on expression of MMP9 or MMP2 in the melanoma tissue. The circulating level of MMP2 declined ∼50% from baseline following rhARSB, declined ∼19% following Pembrolizumab, and had greatest decline (∼58%) by their combination (**Fig.3C**).

**Fig. 3.**
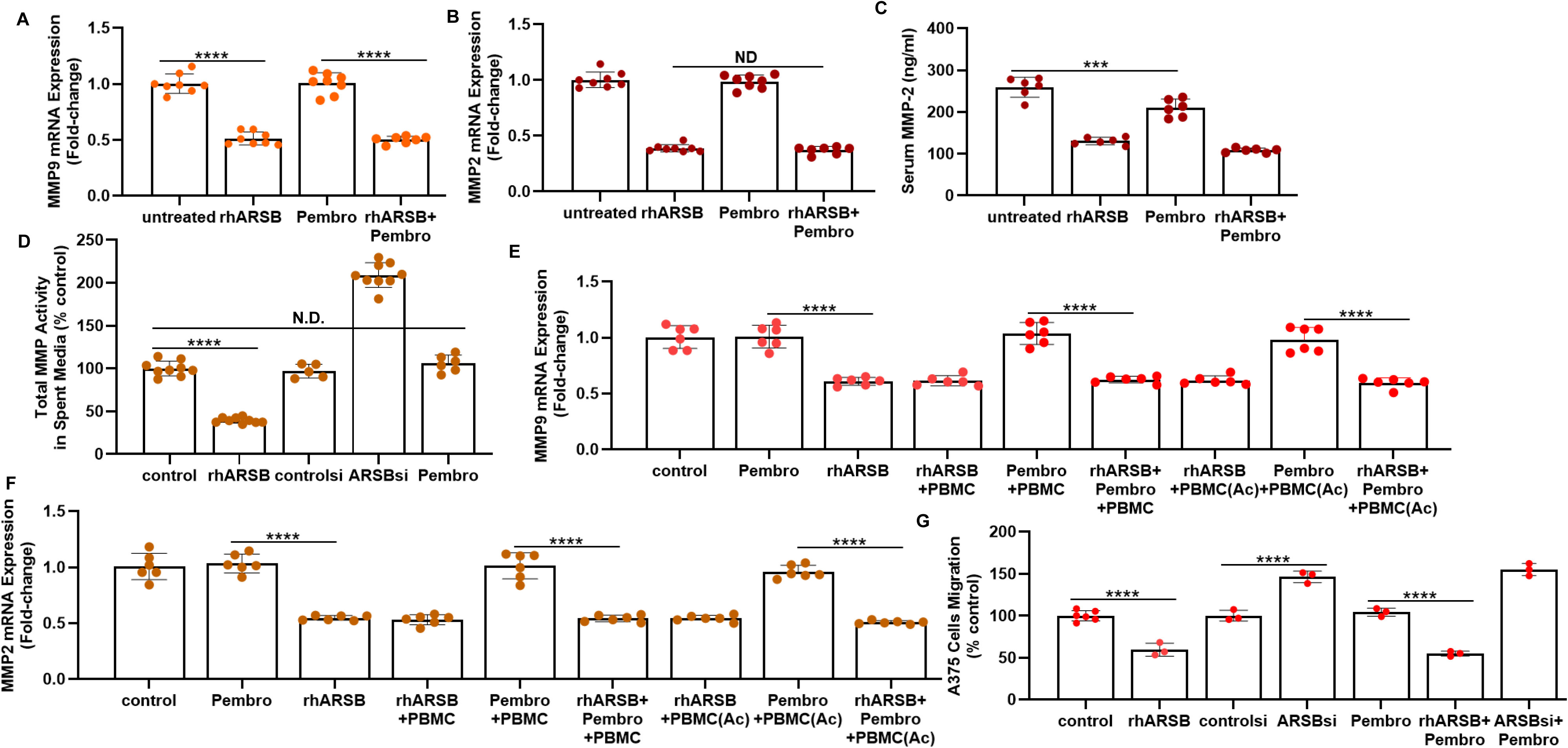
Matrix metalloproteinases following rhARSB and Pembrolizumab in B16F10 pulmonary metastatic melanomas and in A375 cells. **A,B.** mRNA expression of MMP9 and MMP2 declined following rhARSB and was unchanged by Pembrolizumab in the pulmonary melanomas. **C.** Serum MMP2 declined more following rhARSB than following Pembroliziumab and was further reduced by combined treatment in the metastatic pulmonary melanomas. **D.** Total MMP activity in spent media of A375 cells declined following rhARSB increased following ARSB siRNA, but was unaffected by Pembrolizumab. **E,F.** MMP9 and MMP29 mRNA expression in A375 cells was unaffected by Pembrolizumab, either alone or in combination with unactivated or activated PBMC. In contrast, rhARSB significantly reduced their expression. **G.** Migration of A375 cells declined following rhARSB, increased following ARSB silencing, and was unaffected by Pembrolizumab. [Ac=activated; ARSB=arylsulfatase B=N-acetylgalactosamine-4-sulfatase; MMP-matrix metalloproteinase; PBMC=peripheral blood mononuclear cells; Pembro=Pembrolizumab; rh=recombinant human; si=siRNA]

In A375 melanoma cells, MMP activity was reduced by rhARSB, increased by ARSB siRNA, and unchanged by Pembrolizumab (**Fig.3D**). Consistent with these effects on activity, MM9 (**Fig.3E**) and MMP2 (**Fig.3F**) expression declined following rhARSB and was unchanged by Pembrolizumab. The addition of activated or unactivated PBMC had no impact on the MMP expression. Migration of the A375 cells declined following rhARSB, increased when ARSB was silenced, and was unaffected by Pembrolizumab (**Fig.3G**).

The impact of ARSB on MMP expression is mediated by a signaling pathway attributable to effects of decline in ARSB activity on chondroitin 4-sulfation, leading to increased binding of chondroitin 4-sulfate (C4S) with SHP2, the ubiquitous non-receptor tyrosine phosphatase, as previously detailed [3–6]. These mechanisms are presented in A375 cells (**Supplementary Fig. 2A-H**). Also, in normal human melanocytes and B16F10 cells, ARSB siRNA increased MMP and rhARSB reduced MMP expression (**Supplementary Fig.2I, 2J**).

Treatment by rhARSB consistently shows decline in MMP expression and decline in A375 cell migration. These effects may contribute to the *in vivo* effectiveness of rhARSB treatment and suppress metastases. The combination of rhARSB may offer an *in vivo* advantage, since Pembrolizumab had no impact on MMP expression or melanoma cell migration.

### Increases in IL-10 and MCP1 and declines in KC and IL-6 following rhARSB, Pembrolizumab, and their combination in syngeneic mouse model

In the B16F10 metastatic pulmonary melanomas, combined treatment by rhARSB and Pembrolizumab produced greater increases in MCP1 and IL-10 mRNA expression than either agent alone (**Figs.4A,4B**). In contrast, IL-17α was increased by Pembrolizumab, but ARSB had no effect (Fig.4C). Combined treatment produced greater declines in KC, the mouse analog of IL-8, and IL-6 than either agent alone (**Figs.4D,4E**). Overall, there was decline in TNFα due to the inhibitory effect of rhARSB, but, in contrast, Pembrolizumab alone increased TNF-α (Fig.4F). Declines in bFGF, VEGF, and EGF followed rhARSB treatment, and Pembrolizumab had no impact on these declines (**Figs.4G,4H,4I**). The impact of ARSB silencing by siRNA and exogenous rhARSB (1 ng/ml x 24 h) on cytokine expression in A375 cells in a cytokine array showed inverse effects of rhARSB (declines) and ARSB siRNA (increases) on TNF-α, VEGF, IL-6, and FGF-β and was confirmed by mRNA data (**Supplementary Fig.3A-E)**. IL-10 was increased by rhARSB, but unaffected by ARSB siRNA. IL-8 expression mRNA declined following rhARSB and increased following ARSB silencing.

**Fig. 4.**
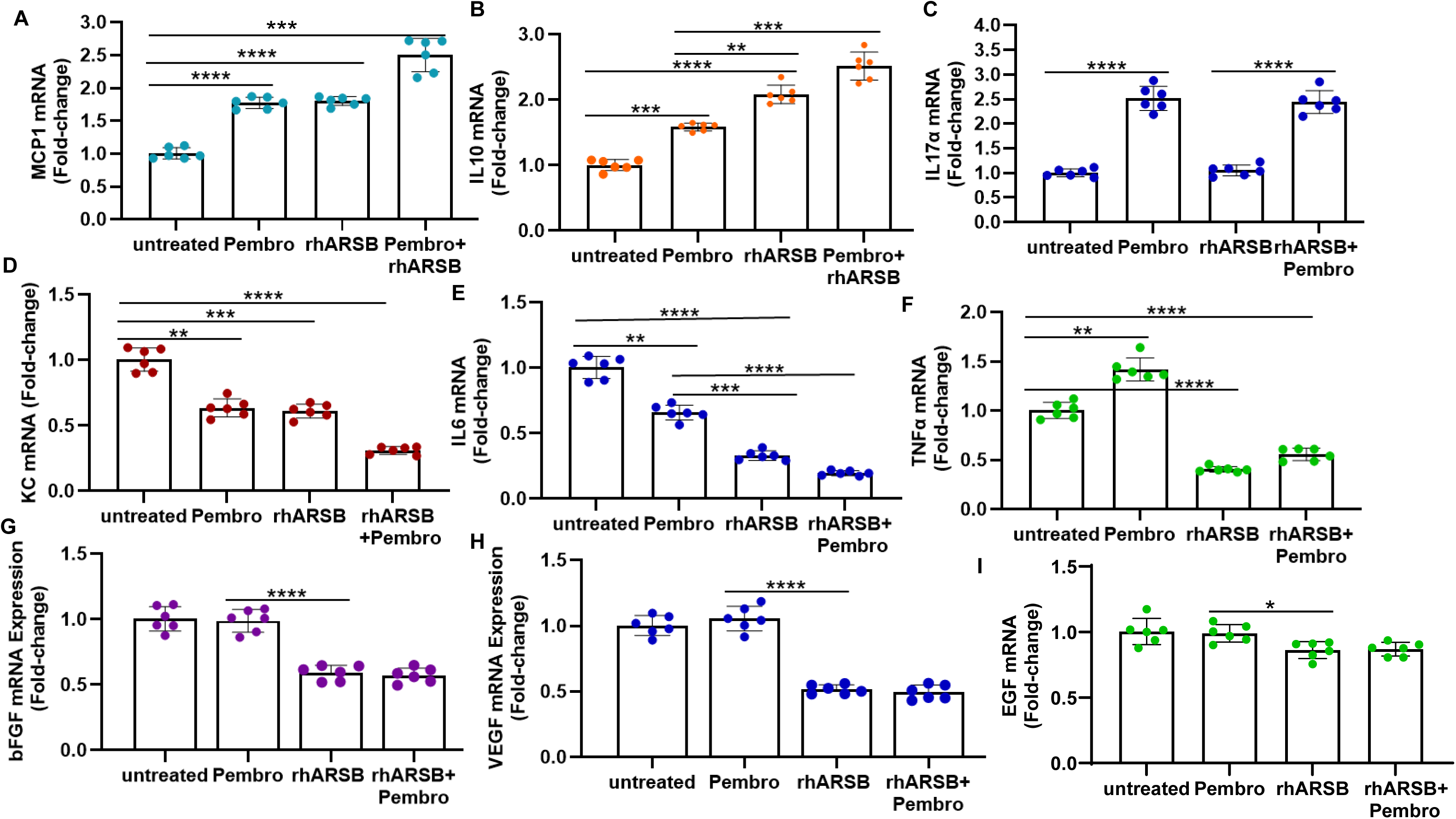
Expression of cytokines/chemokines in B16F10 melanoma lung tissue following rhARSB and Pembrolizumab. **A,B.** In B16F10 pulmonary melanoma tissue, mRNA expression of MCP1 and IL-10 increased following treatment by Pembrolizumab or rhARSB, and the increase was enhanced by their combination. C. In contrast, IL-17α expression was increased by Pembrolizumab, but not by rhARSB. **D,E.** KC, the mouse analog of IL-8, and IL-6 were reduced by both rhARSB and Pembrolizumab and further reduced by their combination. F. In contrast, TNFα was increased by Pembrolizumab and reduced by rhARSB, with overall reduction by their combination. **G,H,I.** FGF2, VEGF, and EGF were reduced by ARSB, but not by Pembrolizumab, and the combination of ARSB and Pembrolizumab did not lead to additional decline. [ARSB=arylsulfatase B=N-acetylgalactosamine-4-sulfatase; EGF=epidermal growth factor; bFGF=basic fibroblast growth factor; IL=interleukin; KC=CXCL1=keratinocyte-derived cytokine; rh=recombinant human; MCP1=CCL2= monocyte chemoattractant protein-1; Pembro=Pembrolizumab; VEGF=vascular endothelial growth factor]

The combined effects of rhARSB and Pembrolizumab on several cytokines reflect interactions by tumor cells and by infiltrating immune cells. The complexity of the tumor microenvironment is apparent with combined anti-inflammatory effects of rhARSB and Pembrolizumab on KC (IL-6), IL-8, and IL10, but different effects on IL-17α and TNF-α.

### Impact of rhARSB and chondroitin 4-sulfate - IL-8 interaction on polymorphonuclear leukocyte (PMN) invasion and apoptosis

PMN invasion toward A375 cells was measured in two-cell layer culture in which A375 cells were plated on the bottom surface of wells and calcein-labeled neutrophils were introduced into the transwell filters on top. PMN passage through the filter was quantified by detection of calcein fluorescence in the lower compartment. Following rhARSB treatment of A375 cells or by addition of exogenous rhIL-8 (**Fig.5A**), PMN invasion increased, since IL-8 in the spent media was increased (Fig.5C). Inversely, silencing ARSB in the A375 cells (**Fig.5A**) reduced the PMN invasion, since more IL-8 was bound to the A375 cells (Fig. 5D). Pembrolizumab alone had no impact on PMN migration, whereas IL-8 in the bottom well increased PMN invasion (**Fig.5B**). Total IL-8 declined following rhARSB and increased when ARSB was silenced (Fig.5E).

**Fig. 5.**
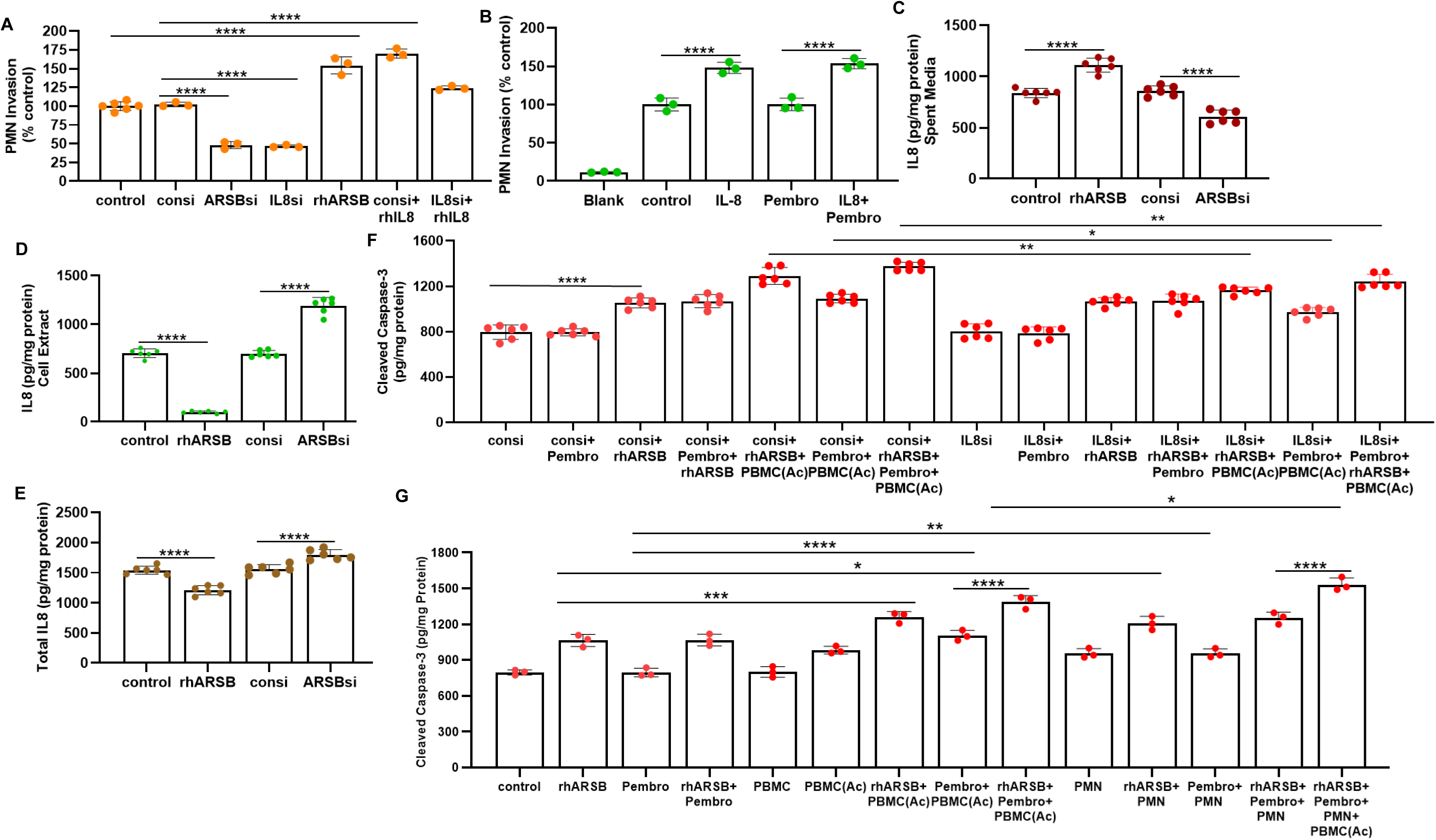
Effects of rhARSB and Pembrolizumab on polymorphonuclear (PMN) leukocyte invasion and apoptosis in A375 cells. **A.** PMN invasion to the A375 cells was increased by rhARSB, reduced by ARSB siRNA, reduced by IL-8 silencing and increased by addition of rhIL-8. These effects are attributable to changes inIL-8 in the spent media. PMN migration from the filter on top to the bottom well was increased by rhIL-8 in the bottom well, but unaffected by Pembrolizumab. **C.** IL-8 in the spent media of A375 cells following rhARSB increased and declined when ARSB was silenced. **D.** In contrast, IL-8 in the cell extract declined following rhARSB and increased following ARSB siRNA. **E.** Total IL-8 in the A375 cells declined following rhARSB and increased following ARSB knockdown. **F.** When IL-8 was silenced in the A375 cells, the effect of rhARSB + PBMC(Ac) and of Pembro + PBMC(Ac) and their combination [rhARSB+Pembro+PBMC(Ac)] on cleaved caspase-3 was reduced, consistent with an effect of IL-8 on PBMC(Ac)-mediated apoptosis. **G.** When PMN were added to the combination of rhARSB+Pembrolizumab+PBMC, the cleaved caspase-3 was further increased in the A375 cells. [Ac=activated, consi=control siRNA; IL=interleukin; rh=recombinant human; N.D.=no difference; Pembro=Pembrolizumab; PBMC=peripheral blood mononuclear cells; PMN=polymorphonuclear leukocytes; si=siRNA]

The effects on PMN invasion are attributable to increased binding of IL-8 to the more highly sulfated chondroitin 4-sulfate of the A375 cells which is present when ARSB activity is less (**Supplementary Figs.4A,4B**). In contrast, chondroitin 6-sulfate does not bind with IL-8 (**Supplementary Fig. 4D**).

When A375 cells were treated by rhARSB, Pembrolizumab and activated PBMC, the effect of activated PBMC on cleaved caspase-3 was significantly reduced by knockdown of IL-8 (**Fig.5F**). The increase in cleaved caspase-3 following rhARSB, Pembrolizumab and activated PBMC was further increased by the addition of PMN for 1 h (**Fig.5F**). These findings indicate that PMN invasion to human melanoma cells *in vitro* is reduced when IL-8 or ARSB is silenced and increases upon addition of rhARSB or rhIL-8 to the surrounding media. *In vivo*, PMN adherence to melanoma cells may be increased when C4S is increased and IL-8 binds more. These perturbations demonstrate the effects of chondroitin 4-sulfate binding with IL-8, which is increased when ARSB is silenced.

## Discussion

Metastatic malignant melanoma remains a devastating disease, and advances in treatment options have the potential to improve quality of life and survival for thousands of patients. With this objective, the current studies have addressed the potential benefits of treatment by rhARSB, checkpoint inhibitor (Pembrolizumab), and their combination in human cell-based and murine *in vivo* studies. The combined effects of rhARSB, acting directly on the melanoma cells, and Pembrolizumab, acting on the infiltrating lymphocytes, suggest the potential for synergism, which may be of significant clinical benefit. Enhanced apoptosis mediated by different BCL2 regulatory mechanisms, decline in matrix metalloproteinases, tumor cell invasiveness and viability, and effects on cytokine expression are all anticipated to inhibit melanoma progression. This report showed no advantage in reduction of pulmonary B16F10 syngeneic metastatic melanomas, by the combination of rhARSB and the human anti-PD-1 Pembrolizumab. However, in this report, the effects on apoptosis, viability, invasiveness, and cytokine expression suggest that the application of combined dosing regimens of rhARSB and checkpoint inhibitor has the potential to enhance the therapeutic benefit.

Interest in the role of chondroitin 4-sulfate (C4S; chondroitin sulfate A; CSA) as a tumor-agnostic, oncofetal antigen reflects the underlying significance of ARSB, since ARSB is the specific chondroitin sulfatase enzyme that removes 4-sulfate groups from the non-reducing end of C4S and is required for the degradation of C4S, as evident in MPS VI [1,2,17–22]. The malarial protein VAR2CSA of plasmodial-infected erythrocytes attaches to the more highly C4S present in the placental vasculature and in small extracellular vesicles associated with adenocarcinoma [22]. These associations are consistent with the impact of ARSB on chondroitin 4-sulfation and the reported effect of the anti-malarial drug chloroquine on the inhibition of ARSB activity [36].

Investigations have shown that activity and/or expression of ARSB is regulated by several potential conditions or exposures [3,12, 32–37]. These include: decline in the cystic fibrosis transmembrane regulator (CFTR) in cystic fibrosis, high chloride, high phosphate, low oxygen, chloroquine, ethanol, carrageenan, estrogen, dihydrotestosterone, and COVID-19. Experiments have indicated that expression of ARSB may be influenced by Rb activation, as mediated by E2F1, phospho-p38α, and cyclin-induced phosphorylation of Rb [37]. Also, decline in ARSB function, both in MPS VI and in human hepatic cells and tissue from ARSB-null mice, is associated with mitochondrial abnormalities [38,39]. These include the presence of unusual inclusions and the disruption of normal mitochondrial function, consistent with impaired aerobic metabolism. The overlap between effects of decline in ARSB function and of hypoxia is consistent with an oxygen requirement for the post-translational modification of ARSB by the formylglycine modifying enzyme (FGE), encoded by SUMF (sulfatase modifying factor) [40,41]. Also, decline in ARSB gene expression was reported in association with unresponsiveness of patients with moderate COPD to oxygen therapy [42].

Several signaling mechanisms by which ARSB independently can affect progression of melanoma and potentially other malignancies have been published [10–15]. These mechanisms are predominantly dependent on ARSB-mediated changes in chondroitin 4-sulfation and the consequent impact on binding with critical signaling molecules, including galectin-3 and SHP2. When ARSB activity is reduced, chondroitin 4-sulfation is increased, and binding of galectin-3 with C4S is reduced. This increases the availability of galectin-3 to influence transcriptional events by enhanced effects of transcription factors, including AP-1 and Sp1. We have shown that increase in free galectin-3 following decline in ARSB function enables increased expression of HIF-1α, versican, CSPG4, and Wnt9A, and PD-L1 by an HDAC3-mediated epigenetic mechanism [3,5,6,10,11, 12,15].

In contrast to reduced binding of galectin-3 with more highly sulfated C4S, the ubiquitous non-membrane tyrosine phosphatase SHP2 (PTPN11), binds more with C4S when ARSB activity is reduced. Signaling effects when SHP2 is bound more with C4S involve extensive persistent phosphorylations with sustained activation of important, diverse mediators. Ser-Thr phosphorylations are influenced through effects on Tyr-Thr phosphorylation sites on phospho-p38, ERK1/2, and JNK. These changes affect transcription factors, including c-Myc, MITF, c-jun, and c-fos and expression of MMP2, MMP9, GPNMB, EGFR, and DNMT with subsequent impact on methylation of the DKK3 promoter and activation of Wnt/β-catenin signaling [3,5,6,13–16].

Additional impact of decline or increase in ARSB activity is attributable to altered binding of more or less sulfated C4S and interactions with other molecules. We have reported increased binding of IL-8, BMP4, and kininogen with C4S when ARSB is reduced [15,34,43]. Increased binding of IL-8 with C4S may affect the attraction or adherence of neutrophils or circulating mononuclear cells to malignant cells. The increased migration of neutrophils toward the A375 cell media in the bottom of the transwells is consistent with increased IL-8 secretion into the media and less IL-8 binding with cell-bound C4S of the A375 cells, when the cell-bound C4S is reduced by rhARSB treatment (**Fig.5, Supplementary Fig.4**). Potentially, reduced binding of PMNs to IL-8 on the surface of melanoma cells following rhARSB and decline in C4S may permit increased access of cytotoxic T-cells to the malignant cells and enhance CD8+-mediated cytotoxicity.

Overall, the studies in this report are evidence of how combined treatments, one directed at enhancing the cytotoxic T-cell response in the tumor microenvironment, and the other at modulating intracellular signaling events and expression of critical signaling molecules in melanoma cells, may together lead to more effective treatment of malignant melanoma and improve survival. Future work will clarify the optimal regimens for combined use of rhARSB and checkpoint inhibitors, anticipating that prior use of rhARSB will reduce the expression of PD-L1 and other potential ligands, enabling enhanced effectiveness of infiltrating immune cells.

## Data Availability

All data produced in the present study are available upon reasonable request to the authors

## Supplementary Figure Legends

**Supplementary Fig.1.**
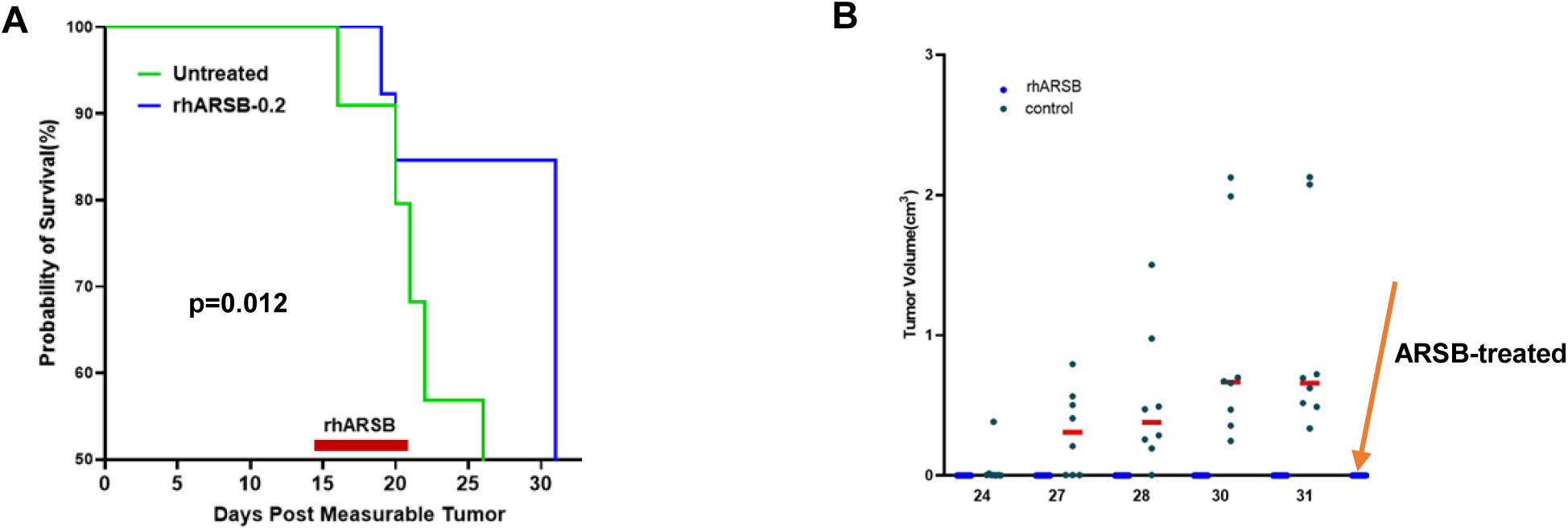
ARSB inhibits progression of subcutaneous B16F10 tumors and development in humanized mice with A375 tumors. **A.** Treatment by rhARSB (0.2 mg/kg in ∼40 µl saline) around the rim of B16F10 subcutaneous tumors in C57BL/6J mice was undertaken for 7 consecutive days in 7 female mice with measureable right flank melanomas. Previous work had treated with rhARSB preemptively on a fixed schedule, prior to the presence of measureable tumors. Initial procedures for tumor inoculation were similar to previous methods [5]. Survival was compared to 8 untreated mice which were treated with similar volume of normal saline at similar time points. Tumor volume was reduced by ARSB treatment at 6 of 7 time points. Survival was increased in the treated mice, from mean of 26 days in the controls to 31 days in the treated mice (p=0.012, log-rank test). **B.** Immune-deficient female mice NSG^TM^ humanized mice (NOD.Cg-*Prkdc*^scid^ Il2rg^tm1Wjl^SzJ; strain #005557, Jackson Labs, Bar Harbor, ME) were engrafted with human PBMC and subsequently inoculated with 200,000 A375 human melanoma cells subcutaneously in the right flank. Treatment was rhARSB (0.2 mg/kg SQ at a concentration of 50 µg/ml saline) on days 2, 7, 14, 21, and 28 following tumor inoculation. Seven mice were treated with rhARSB, and 8 controls were treated with similar volume of saline on the same schedule. Mice were euthanized on day 31, when two of the untreated mice reached endpoint for tumor volume. All of the untreated mice had measureable tumors. In contrast, no tumors were palpable or visible in the treated mice.

**Supplementary Fig.2.**
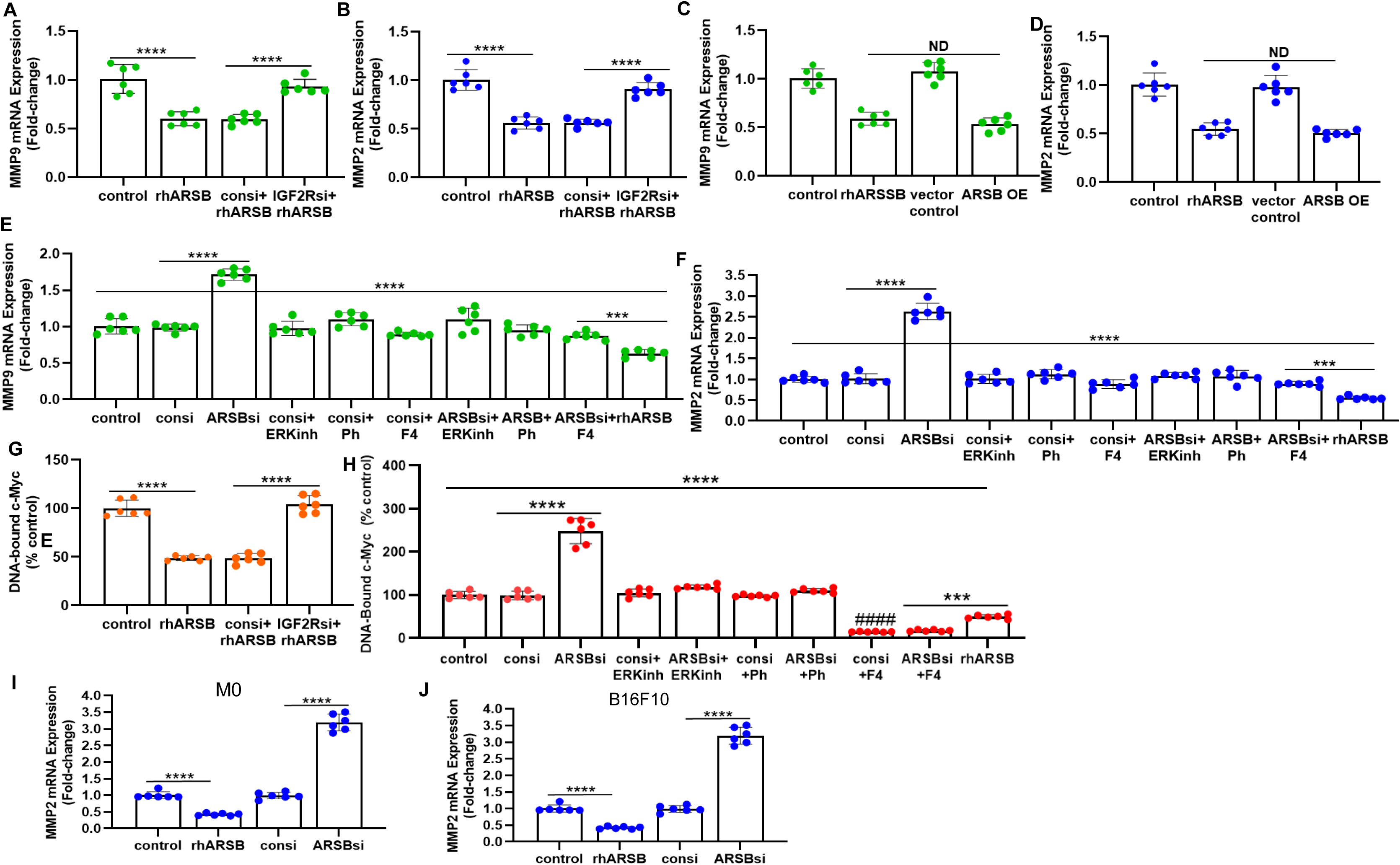
ARSB-induced effects on MMP expression in A375 cells by effects on IGF2R and c-Myc. **A,B**. In A375 cells, the inhibitory effect of rhARSB on MMP9 and MMP2 expression was blocked by knockdown of the IGF2R (insulin-like growth factor 2 receptor), the mannose 6-phosphate receptor which enables uptake of rhARSB [4,13]. **C,D**. Overexpression of ARSB by specific plasmid had a similar effect on MMP9 and MMP2 expression as treatment by rhARSB. Overexpression was performed using ARSB plasmid in pCMV6-XL4 vector (OriGene, Rockville, MD) and introduced in A375 cells by transient transfection using 2 μg of the plasmid and Lipofectamine^TM^ 2000 (Invitrogen, Thermo Fisher Scientific). Previously, controls included untransfected cells and cells transfected with empty vector control (OriGene). Media were exchanged after 6 h. Cells were incubated in a humidified, 37°C 5% CO_2_ environment and harvested at 24 h after transfection. Effectiveness of transfection was determined by measurements of ARSB activity before and after transfection. ARSB activity was measured as previously reported [3,6]. **E,F**. Increases in MMP9 and MMP2 mRNA were inhibited by ERK activation inhibitor peptide 1, cell-permeable (ERKinh), p38-MAPK inhibitor (Ph), and c-Myc/Max inhibitor (F4; thiazolidinone) which blocks c-Myc-Max dimerization. Prior experiments in subcutaneous B16F10 melanomas and in cultured melanoma cells showed that ARSB affected the expression of MMP2 and MMP9 through SHP-2-mediated effects on phospho-ERK1/2 and subsequent effects on DNA binding of c-Myc [5,6]. Similar effects are shown in the A375 cells. **G.** DNA-bound c-Myc was inhibited by treatment with rhARSB in A375 cells. Knockdown of IGF2R, which is required for cellular effects of exogenous rhARSB, inhibited this response [4,10]. **H.**. ERK activation inhibitor peptide 1, cell-permeable (ERKinh), p38-MAPK inhibitor (Ph), and c-Myc/Max inhibitor (F4) blocked the increase in nuclear-bound c-Myc. ARSB siRNA increased DNA-bound c-Myc, and this increase was inhibited by ERK inhibitor, p38-MAPK inhibitor, and c-Myc/Max inhibitor (cell-permeable thiazolidinone, labelled F4), which completely suppressed the effect of ARSB silencing on DNA-bound c-Myc. **H.** ARSB knockdown increased MMP2 expression in normal human melanocytes. The normal melanocytes were grown and treated as previously described [6,10]. Primary normal melanocytes were cultured in Airway Epithelial Cell Basal Medium (ATCC, Manassas, VA) with melanocyte growth kit (ATCC) and maintained at 37°C in a humidified, 5% CO2 environment with media exchange every 3 days. Confluent cells were harvested by trypsin for primary cells (ATCC) and sub-cultured. **I.** Consistent with other findings, rhARSB significantly reduced mRNA expression of MMP2 and MMP9 (not shown) and ARSB siRNA significantly increased MMP2 and MMP9 (not shown) expression in B16F10 mouse melanoma cells. There was greater increase of MMP2 than MMP9 by ARSB knockdown. B16F10 mouse melanoma cells were purchased (ATCC CRL-6475) and cultured in DMEM supplemented with 10% FBS and 1% penicillin-streptomycin. Cells were screened for pathogens by IDEXX BioAnalytics (Columbia, MO). Cells were maintained at 37°C in a humidified, 5% CO2 environment with media exchange every 2 days, and confluent cells in T-25 flasks were harvested by EDTA-trypsin, and sub-cultured.

**Supplementary Fig.3.**
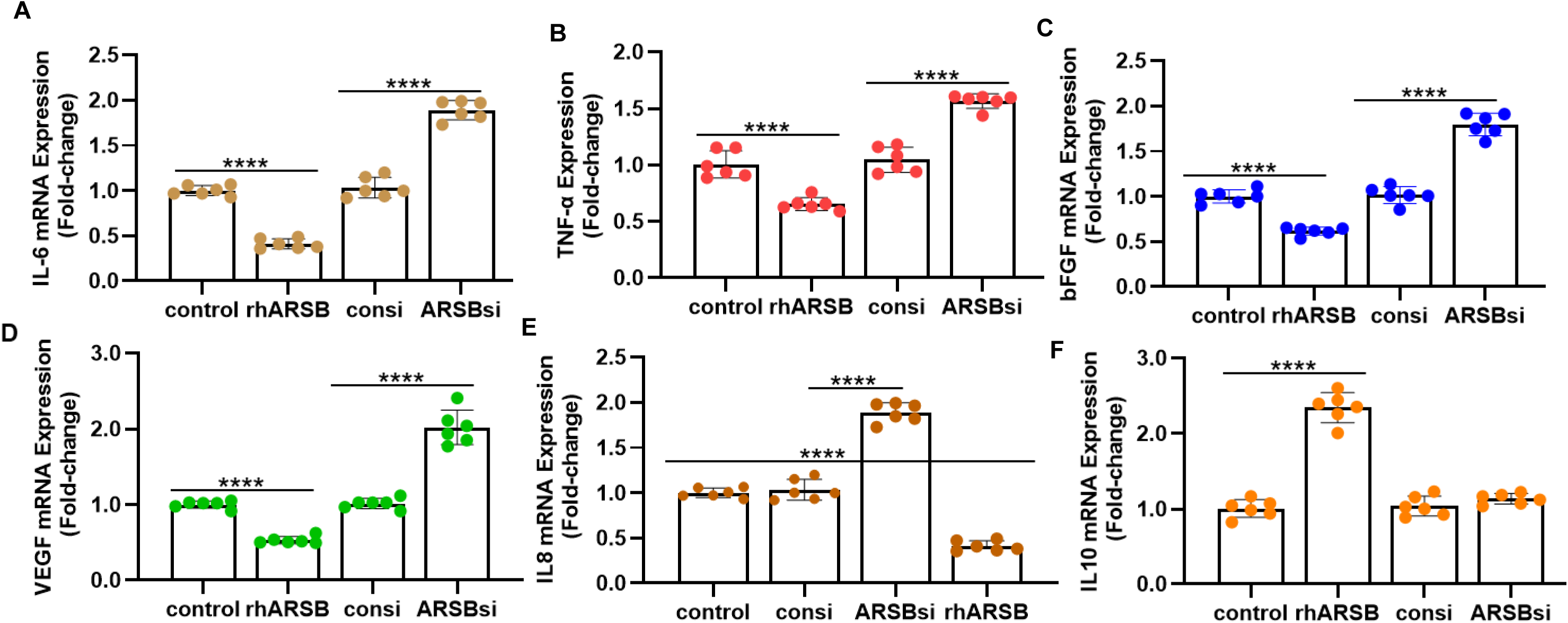
Cytokines/Chemokines following ARSB siRNA and rhARSB in A375 cells. **A.B, C,D,E**. mRNA expression data show inverse effects of rhARSB (declines) and ARSB siRNA (increases) on IL-6, TNF-α, bFGF, VEGF, and IL-8. These results are similar to findings from cytokine array. F. IL-10 is increased by rhARSB and unaffected by ARSB siRNA.

**Supplementary Fig. 4.**
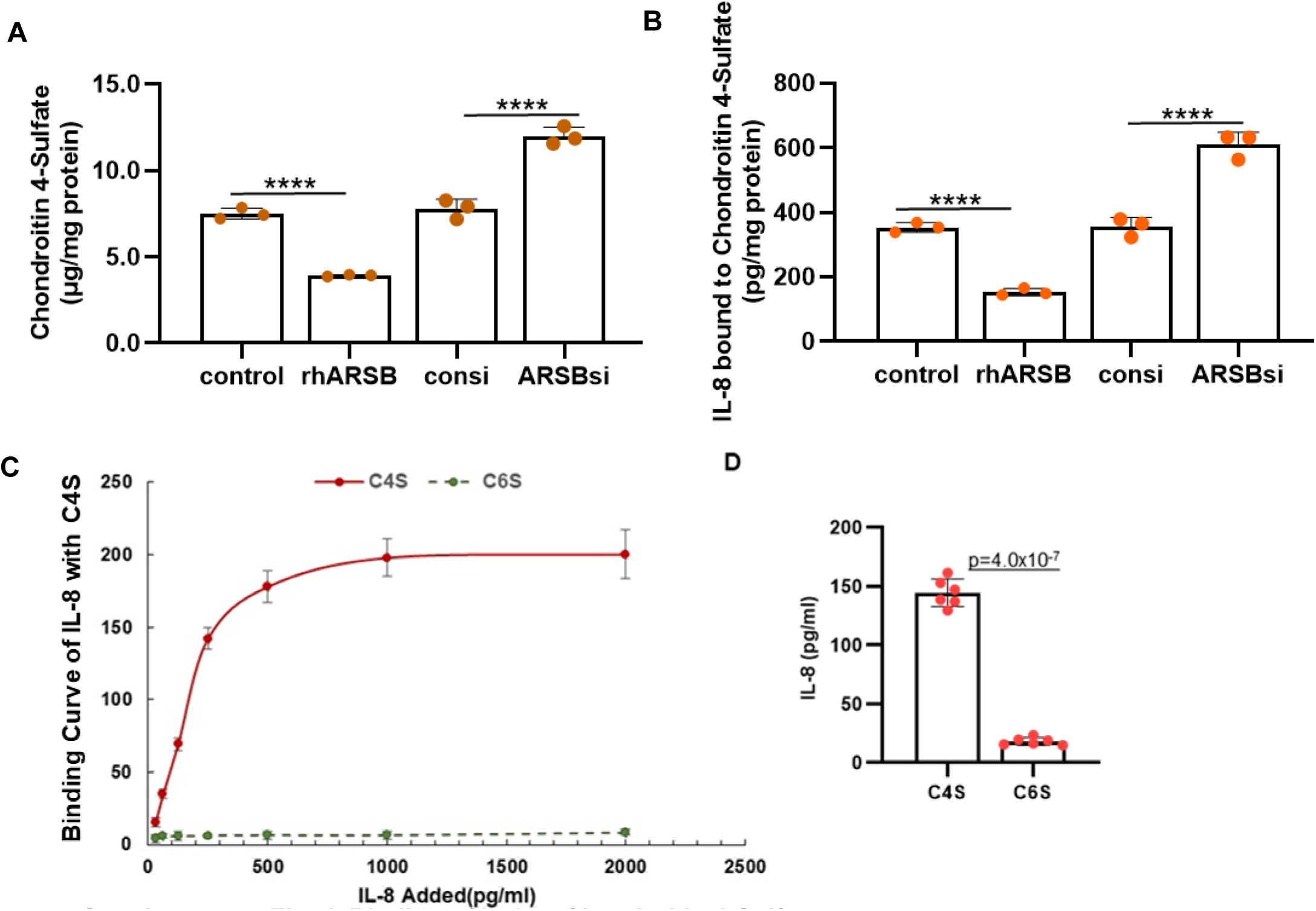
Binding of IL-8 with Chondroitin 4-Sulfate. **A.** Chondroitin 4-sulfate content in A375 human melanoma cells was reduced by rhARSB (1 ng/ml x 24h) and increased when ARSB was silenced by siRNA. **B.** When rh IL-8 was added to wells of a 24-well plate coated with A375 cells, the amount of bound IL-8 declined when cells were treated with rhARSB and increased when ARSB was silenced, indicating that IL-8 binds more with more sulfated C4S present when ARSB activity is reduced. IL-8 in the cell extracts was measured by ELISA IL-8 (DY208, R&D Systems). **C**. In an *in vitro* binding experiment, wells of a 24-well plate were coated with either C4S or C6S antibody (5 µg/ml) and C4S or C6S (100 µl; 2.5 µg/ml) was added. Subsequently, rhIL-8 at varying concentrations was added, biotinylated IL-8 antibody was added, and the extent of IL-8 binding detected by ELISA, as detailed in the Methods. Maximum binding of IL-8 with C4S was ∼200 pg/ml. Virtually no IL-8 bound to chondroitin 6-sulfate. **D**. When 250 pg/ml of IL-8 was added to wells coated with either C4S or C6S, 144.6 ± 11.7 pg/ml of IL-8 bound to C4S and 18.3 ± 3.2 pg/ml of IL-8 bound to C6S.

**Supplementary Fig. 5.**
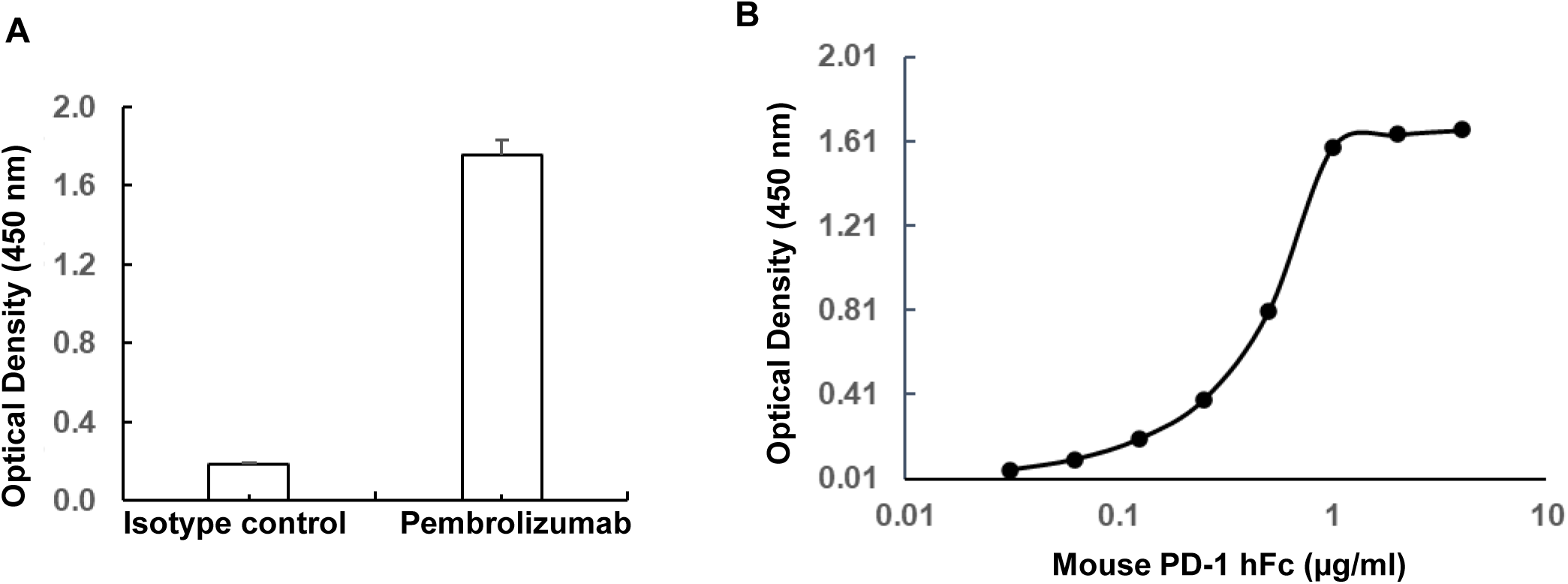
Binding of mouse PD-1 with Pembrolizumab. **A.** IGG4-kappa isotype control binding to mouse recombinant human Fc-tagged PD-1 was minimal. In contrast to binding to Pembrolizumab in coated wells. **B.** Binding to Pembrolizumab increased with increasing concentration of mouse PD-1, peaking at ∼1.61 µg/ml.

